# Systematic Review of Supervised Machine Learning Models in Prediction of Medical Conditions

**DOI:** 10.1101/2022.04.22.22274183

**Authors:** Branimir Ljubic, Martin Pavlovski, Avrum Gillespie, Daniel Rubin, Galen Collier, Zoran Obradovic

**Affiliations:** Temple University, Center for Data Analytics and Biomedical Informatics (DABI), Philadelphia, PA 19122, U.S.A.; Rutgers University, Office of Advanced Research Computing (OARC), Piscataway, NJ 08854, U.S.A.; Lewis Katz School of Medicine, Temple University, Philadelphia PA 19140, U.S.A.

## Abstract

Machine learning (ML) models for analyzing medical data are critical for both accelerating development of novel diagnostic and treatment strategies and improving the accuracy of medical care delivery. Our objective was to comprehensively review supervised ML models for diagnosis or treatment prediction. Publications indexed in PubMed were reviewed to identify articles utilizing supervised predictive ML models in medicine. Articles published between 01/01/2020–01/01/2022 were included in this review. Initially, PubMed was searched using MeSH major terms, and if more extensive search results were needed, a broader search was applied (titles/abstracts).

PubMed indexed 21,268 published articles (MeSH Major topic) describing ML methods implemented in medicine. Of those, 11,726 articles were published within the last 2 years. Most of the published ML models in medicine in the last two years were different types of deep learning models (about 75%). Fifty articles were included in this review.

Almost all categories of disease were subjects of ML predictions. Positive and negative factors in each of the scenarios need to be evaluated before the most optimal ML model is selected. Domain knowledge and collaborations between physicians and ML experts can improve the selection and prediction performance of ML models in medicine and facilitate implementation in clinical practice. Predictive ML models could provide recommendations to recruit suitable patients for clinical trials. Prediction ML models may contribute to development of more effective diagnostic and therapeutic choices, founded on evidence-based medicine. A broad range of methodological approaches have been taken toward this goal, and those approaches are presented here with their various advantages and disadvantages.

**AUTHOR SUMMARY:** Over the last decade, there has been rapid development of Machine learning (ML) methods to analyze Big Data in medicine. ML is aimed to make the computer learn from past experiences and make predictions by recognizing patterns in medical data. We performed a comprehensive systematic literature review of recent publications (last two years), indexed in PubMed/MEDLINE that have described either traditional or deep supervised prediction ML models in medicine. We identified 21,268 articles describing ML implementation in medicine. 11,726 articles were published in the last 2 years. We presented the number of publications describing each of the most often ML methods to show current trends in development of these models. Most of the recently published ML models in medicine were deep learning models. We found that the understanding of disease is likely to lead to more accurate prediction. An important dilemma is the selection of optimal ML models for a specific task, considering amount and type of available data. Domain knowledge and collaborations between physicians and ML experts can improve the prediction performance of ML models, which could help clinicians to select the most effective diagnostic and therapeutic choices available and decrease medical errors.

## INTRODUCTION

Over the last decade, there has been significant growth of the amount of medical data generated by the adoption and integration of electronic health records (EHR).[1] This growth in EHR data coincided with rapid development of ML techniques and computing power to analyze Big Data in medicine, which could contribute to improved medical solutions and better, more efficient healthcare.[2-5] ML is a branch of Artificial intelligence (AI), aimed to make the computer learn from past experiences and make predictions by recognizing patterns in medical data.[5-7] ML can be classified into three categories: unsupervised, supervised, and reinforcement learning (RL). This paper focuses on supervised ML techniques, where a function that maps an input to an output is inferred from labeled training data. The objective of the research was to perform a comprehensive systematic literature review of recent publications that have used either traditional or deep supervised prediction ML models in medicine. We examined whether the method is appropriate for the selected medical prediction task, whether the model is generalizable, and whether it could be used by clinicians to improve the quality of medical care. Supervised prediction ML models are utilized in traditional and deep learning approaches.[5-7] The most frequently used traditional ML models in medicine are: decision trees (DT),[8] random forest (RF),[9] and other ensemble methods,[10-14] single and multi-layer perceptron (MLP),[15.16] Bayesian learning (BL),[17] support vector machines (SVM),[18] k-nearest neighbors (k-NN),[19] linear regression (LR),[20] and logistic regression (LogR).[21] Deep learning models are inspired by biological neural networks, where each layer of the network learns higher order features of the previous layer. Different types of neural networks have been designed, such as: deep neural networks (DNN) including deep belief networks,[22] convolutional neural networks (CNN),[22] recurrent neural networks (RNN - long short-term memory (LSTM) and gated rectified unit (GRU)),[23] etc.

Many ML models have been developed for prediction of diagnosis, or recommendation of the most optimal therapeutic approach. Since this is a rapidly evolving scientific field, we reviewed articles published within the last two years.

## MATERIALS AND METHODS

We conducted a systematic review of articles describing supervised prediction ML models (traditional and deep learning) published within the last two years (01/01/2020 –01/01/2022). The following traditional supervised ML models were included in the review: DT, RF and other ensemble methods, the perceptron, BL models, SVM, k-NN, LR, and LogR. Deep learning models included in the review were: DNN, CNN, and RNN.

We searched PubMed/MEDLINE, as the most relevant source of medical topics, to identify articles describing applications of supervised traditional and deep predictive ML models in medicine. Initially, we searched PubMed using MeSH (Medical Subject Headings) major terms. If we did not find enough literature for the specific ML model, we applied a broader search using terms that appear in titles of publications, or in case of bagging and boosting ensemble models and RNN we used combinations of MeSH and title/abstract searches. This searching approach extracted articles where the reviewed ML model was a major part of the article. The search strategy used keywords indicating “ML model” AND “prediction/detection/classification” AND “medical conditions (therapies, outcomes)”. In the example of deep learning methods, we divided searches into 3 groups: classic DNN, CNN and RNN. To extract general DNN models we used the following search query: (“deep learning”[Mesh] not “CNN” not “RNN” not “LSTM” not “GRU”). For CNN models the query was: (CNN[Title]) OR (convolutional neural networks[Title]). And for RNN model the query was: (RNN[Title/abstract] OR recurrent neural networks[Title] OR LSTM[Title] OR GRU[Title] OR long short term memory[Title] OR gated rectified unit[Title]).

We included articles that presented original research published in English. The search results were sorted according to types of described ML models. For each of the reviewed ML models we selected a representative sample. Priority was given to the newest published research in cases of multiple papers describing similar predictive ML approaches. At least four authors agreed that the article was sufficiently significant to be included in this review. We performed a systematic review of the literature to analyze how useful and meaningful the described predictive ML models are in terms of realistic applicability in medical practice.

## RESULTS

We identified 21,268 articles (MeSH Major topic) describing ML implementation in medicine (Figure 1). 11,726 articles were published in the last 2 years.

**Figure 1.**
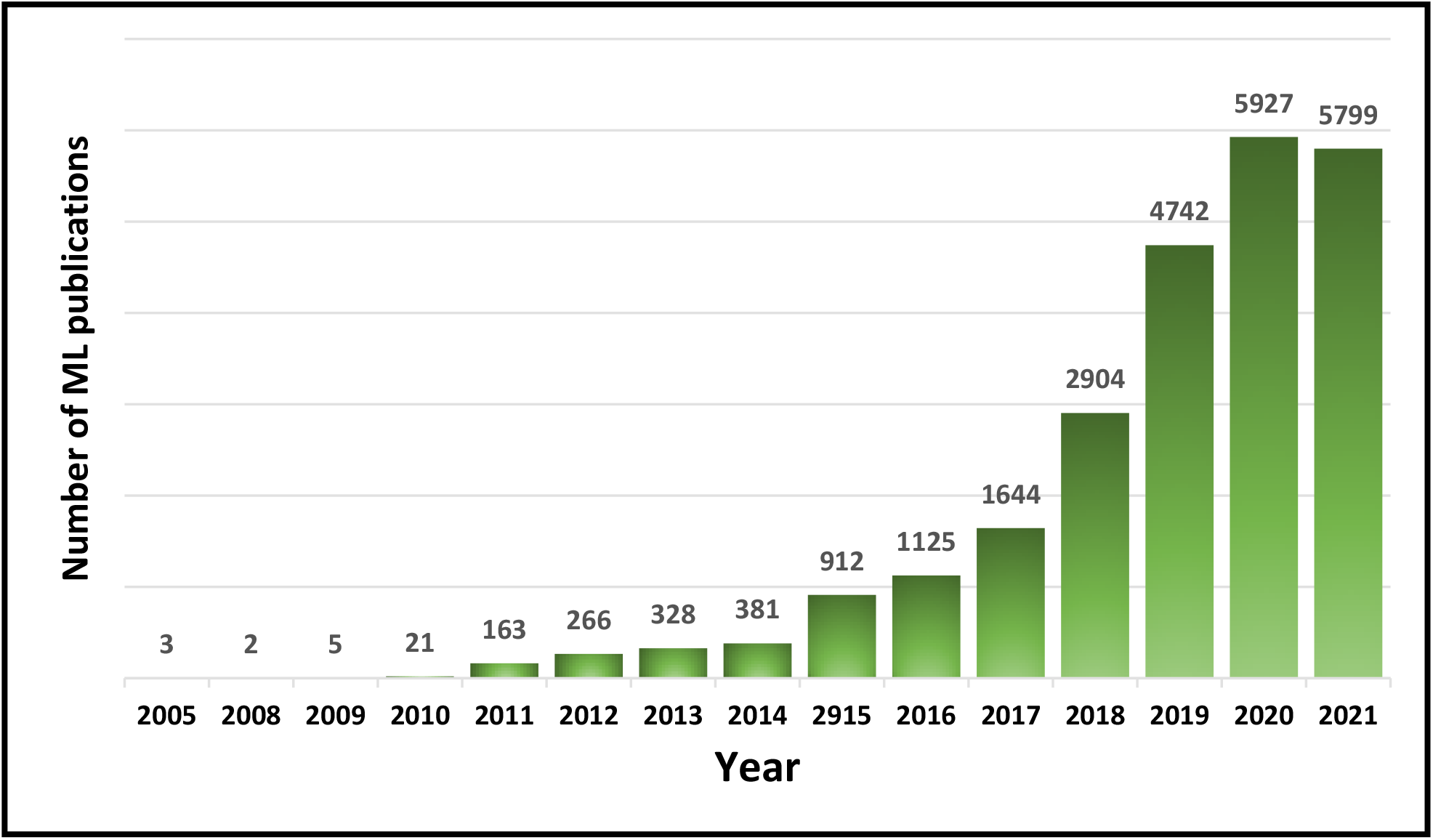
Number of publications describing ML applications in Medicine indexed in PubMed.

Fifty articles were included in the review. In Table 1, we present the method of search for each of the ML models and the number of publications retrieved using that particular search method. Most of the published ML models in medicine in the last two years were different types of deep learning models.

**Table 1.**
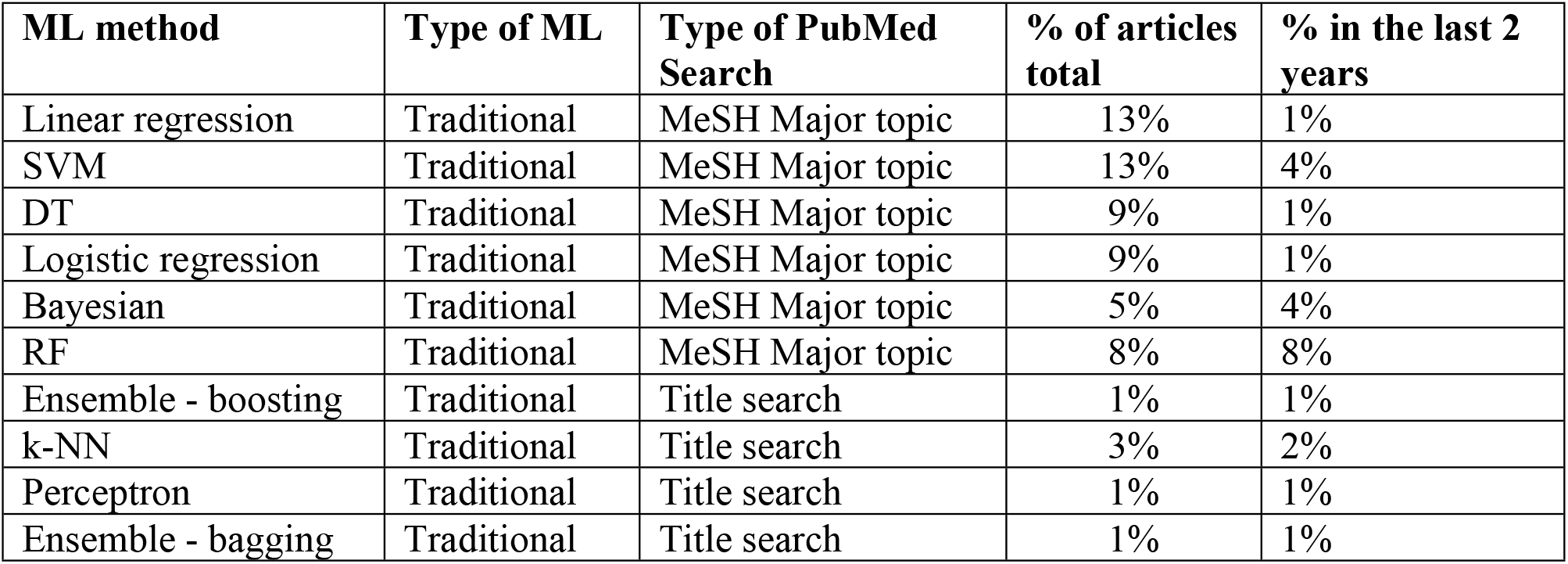
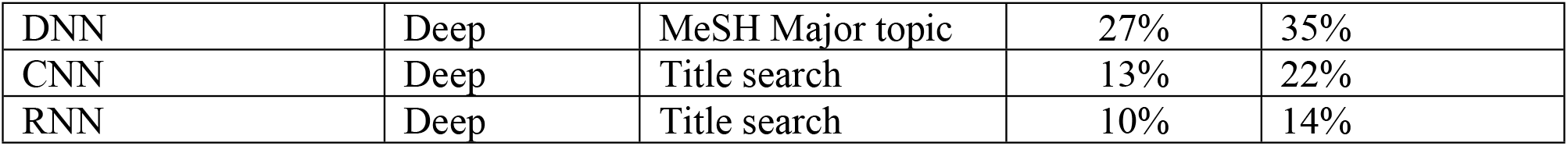
Supervised ML methods applied in prediction of medical conditions. Percentages of articles of the total number of publications by ML method are presented.

| ML method | Type of ML | Type of PubMed Search | % of articles total | % in the last 2 years |
| --- | --- | --- | --- | --- |
| Linear regression | Traditional | MeSH Major topic | 13% | 1% |
| SVM | Traditional | MeSH Major topic | 13% | 4% |
| DT | Traditional | MeSH Major topic | 9% | 1% |
| Logistic regression | Traditional | MeSH Major topic | 9% | 1% |
| Bayesian | Traditional | MeSH Major topic | 5% | 4% |
| RF | Traditional | MeSH Major topic | 8% | 8% |
| Ensemble - boosting | Traditional | Title search | 1% | 1% |
| k-NN | Traditional | Title search | 3% | 2% |
| Perceptron | Traditional | Title search | 1% | 1% |
| Ensemble - bagging | Traditional | Title search | 1% | 1% |
| DNN | Deep | MeSH Major topic | 27% | 35% |
| CNN | Deep | Title search | 13% | 22% |
| RNN | Deep | Title search | 10% | 14% |

### Traditional models

#### Decision trees

are classification methods that adopt a top-down strategy where each node represents a classification question and the branches partition the data into different classes.[8] DT models were developed for prediction of diabetes mellitus type 2 (DM2), and essential hypertension (EH),[24] by creating visually guided classification trees to facilitate the feature selection (four different datasets, sizes 547 – 12,447). The prediction accuracy of DM2 and EH in different scenarios varied between 58% and 87%. DTs predicted coronary artery disease with the accuracy about 91% on a dataset of 303 patients.[25] A DT algorithm was proposed to identify pre-treatment clinical predictors of survival in rectal cancer (100 examples).[26] Predicted accuracy of survival rates were 71-76%. Presented DT models have questionable generalization potential, since most of them were developed on small samples frequently from one hospital.

#### Random Forest

is a ML method that applies many decision trees to predict the outcome.[9] RF models have been constructed to predict hepatotoxicity on a dataset of 346 samples, with the accuracy up to 71%.[27] This may provide a basis for improved safety evaluation in drug discovery and the risk assessment of environmental pollutants. A retrospective review of 559 patients undergoing abdominal hernia was the basis for RF modeling to predict surgical approach and determine the importance of different socioeconomic variables in selecting the type of surgery (area under the receiver operating characteristic (AUROC) ≈0.82).[28] Data were obtained from a single institution, which limits the generalizability of findings. Psychotic and depressive symptom clusters in dementia were predicted using RF on the EHR records of 4,003 patients with dementia (AUROC 0.80).[29] An RF model boosted by the AdaBoost algorithm was utilized to predict the severity of COVID-19 cases and the possible outcome, by using a patient’s geographical, travel, health, and demographic data (3397 patients, accuracy 94%). The model revealed a positive correlation between patients’ genders and deaths, finding that men are more likely to die.[30] All presented RF models need further improvements including potential combination with deep learning ML models to find a relevant application in practice.

#### Ensemble Methods

(boosting, bagging) combine a few weak learning models and turn them into a great learning algorithm.[10-14] The most famous boosting algorithm is AdaBoost. ML models were built to predict the probability of different pairs of drugs and nanoparticles creating drug-decorated nanoparticle (DDNPs) complexes with anti-glioblastoma activity. Forty-one features have been selected for 855,129 drug-nanoparticle complexes. The best model was obtained with the bagging ensemble classifier, based on 20 decision trees with the AUROC of 0.96 and accuracy of 87%. This model could be applied for the screening of nanoparticle-drug complexes in glioblastoma.[31] The bagging ensemble method improves reproducibility of both cortical and subcortical functional parcellation of the human brain neuroimaging (more than 300 samples).[32] AdaBoost was used to differentiate colorectal neoplasia from normal tissue (AUROC up to 0.95 on 64 samples from 16 patients).[33] Both models use imaging data as inputs and need additional research work, including comparison to deep learning models, before their potential application in clinical practice.

#### Perceptron and Multilayer Perceptron

The perceptron is based on a threshold function that learns weights for features and processes one example at a time. It could be used as a single-layer perceptron or as a multilayer perceptron.[15,16] Modeling of the spread of the COVID-19 infection using a MLP was designed on a dataset (20,706 examples) operated by the Johns Hopkins University Center for Systems Science and Engineering (JHU CSSE).[34] This is one of many papers about COVID-19 intended to predict the spread of the infection, but the model lacks generalization ability. MLP was applied in a diagnosis of breast cancer subtypes, using MRI images (704 images) with AUROC of 0.86.[35] The study used imaging data to distinguish between benign and malignant breast lesions. The challenge remains how to effectively incorporate this model into everyday oncology.

#### Bayesian ML

algorithms calculate probabilities for hypotheses. The class having maximum probability is assigned as the most suitable class.[17] A study presented a method for classifying electrocardiogram (ECG) data into four emotional states according to the stress levels using naive Bayes and SVM algorithms with the average accuracy of the stress classification of 97.6%. Ability to quantify the stress signals could facilitate a more effective management of mental state.[36] A Bayesian ML model was designed to estimate the probability of an individual having an oral Human Papilloma Virus (HPV) infection, given Oropharyngeal Squamous Cell Carcinomas (OPSCC) and other covariate information.[37] The model is then inverted using Bayes’ theorem to reverse the probability relationship. The authors analyzed 8,106 OPSCC patients and achieved the AUROC of about 0.7. The Bayesian model could be utilized to identify risk factors in estimating the probability of medical conditions.

#### Support Vector Machine

models apply an optimization problem that attempts to find a separating hyperplane with as large a margin as possible.[18] SVM was trained on 318 samples to distinguish neurodegenerative movement disorders such as Parkinson’s Disease (PD) from healthy subjects, and from other movement disorders (precision ≈81% and recall ≈89%).[38] In this study, DNN and RF were applied to the same task, with DNN achieving the best results. Models were trained on retrospective data at a single site, with high data quality and none of the different classifiers outperformed the others, which are some of limitations of these models. SVM models have also been used for diagnosis of early breast cancer using PET images (116 samples, accuracy up to 85%, AUROC 0.89),[39] and detection of atrial fibrillation (AF) using ECG data (79 AF and 336 non-AF cases, accuracy 97-100%).[40] Both models use images as inputs. These models must be tested on bigger data and compared to deep learning since recent literature show that deep learning models yield better prediction than SVM models on imaging data.

#### The K-Nearest Neighbors

algorithm takes into account k-neighboring points when classifying a data point and assigns the class by finding the most prominent class among the k-nearest data points.[19] Hepatocellular carcinoma (HCC) dataset from the UCI machine learning repository was used to test different algorithms, including k-NN classifier, for feature selection and classification. The best results achieved up to 84% accuracy.[41] The k-NN and SVM classifiers were utilized to determine whether the patients have abnormal or normal respiration, or have bradypnea (slow breathing), or tachypnea (fast breathing). The testing accuracies of the completely built SVM and k-NN classifiers were 96% and 99%, respectively.[42] Constructed models must be tested on more datasets from other institutions to determine reproducibility of the models. K-NN models have not been used very often for medical predictions in the last few years.

#### A Linear Regression

algorithm performs a regression task and predicts a specific value based on an independent variable.[20] A linear model was created to predict the impact of the duration of exposure (number of days) to COVID-19 on mortality rates (more than 270,000 patients).[43] Multiple regression and LR analysis were successfully applied to predict the number of weekly deaths due to COVID-19 in India (606 patients).[44] LR models are good options for time to event type of predictions and for predictions of the exact numbers of patients, or the cost of care.

#### Logistic Regression

uses the logistic function to binary classification and estimates the probability of the event.[21] Multivariate LogR (as well as RF and XGBoost) models were applied for prognosis of mortality risk in patients with COVID-19 (292 patients, AUROC ≈0.95).[45] The model needs to be tested on larger multi-center data. A study investigated the application of LogR and RNN LSTM models in capturing clinical risk factors for outcome prediction of 575 patients with aneurysmal subarachnoid hemorrhage (AUROC 0.89).[46] Since, the LSTM RNN model achieved higher accuracy, it is likely a better choice in this type of study. A logistic regression-based ML prognostic algorithm is implemented in real-time as a clinical decision support (CDS) system to facilitate decision making for patients with suspected COVID-19 in the emergency department (ED). Training data included 1,469 adult patients who tested positive for Severe Acute Respiratory Syndrome (SARS) within 14 days of acute care. The algorithm performed well with an AUROC of 0.85.[47] A LogR based ML-enabled CDS can be developed, validated, and implemented with high performance across multiple hospitals while being equitable and maintaining performance in real-time validation.

### Deep learning

#### Deep Neural Networks

DNN is a multilayer neural network with an input layer, hidden layers, and an output layer. DNN learns weights so the output from the network correctly classifies the example. The back propagation algorithm is a standard approach to train DNNs.[22] A compartmental model enhanced with deep learning methodology predicted the dynamics of the COVID-19 epidemic in the U.S. using the JHU CSSE data repository.[48] The model predicted the number of active cases between 3.2-3.3 million on August 16-18, 2020. The actual number of infected cases on August 16-18, 2020, was about 2.5 million (CDC data), so the model was not accurate. DNN methods were designed, to predict the dynamics of the COVID-19 pandemic outbreak on JHU CSSE data and Korea Centers for Disease Control and Prevention data.[49] Predictions of different aspects of COVID-19 epidemic are popular, but considering the current state of the pandemic, it is difficult to confirm that these models work accurately in reality. A DNN was designed for prediction of behavior of engineered RNA elements capable of detecting small molecules, proteins, and nucleic acids.[50] This work shows that DNN approaches could be used for predictions in RNA synthetic biology, but more data are needed for the training of DNNs, as well as improvement of DNN architectures. A DNN performed an automatic diagnosis of the 12-lead ECG recordings and outperformed cardiology residents in recognizing six types of abnormalities, with F1 scores above 80% and specificity over 99%.[51] Additional studies could test weather DNN effectively diagnose different ECG abnormalities, including myocardial infarction. DNN, RF, and a simple statistical test were used to predict COVID-19 infections from full blood counts only (598 samples), without knowing the history of the patients (accuracy up to 91%).[52] It appears that these DNN models are more of a theoretical work at this stage of development, without evident clinical implementation. A deep ML model was developed for automatic detection of brain metastases that uses contrast-enhanced and non-enhanced CT images as inputs. The dataset contained CT scans of 116 patients with brain metastases (total of 659 metastases). Single-shot detector (SSD) ML models were constructed with a feature fusion module. The sensitivity was 88.7% for the model that used both contrast-enhanced and non-enhanced CT images (the CE + NECT model) and 87.6% for the model that used only contrast-enhanced CT images (the CECT model).[53] The model is a contribution to imaging-based diagnostics, and it needs further testing on larger datasets to improve generalization. Another study proposed a novel deep learning architecture involving combinations of CNN layers and RNN layers that can be used to perform segmentation and classification of five cardiac rhythms based on ECG recordings. The algorithm is developed in a sequence to sequence setting where the input is a sequence of five second ECG signal sliding windows and the output is a sequence of cardiac rhythm labels. Experimental result shows this approach can achieve an average F1 scores of 0.89.[54]

#### Convolutional Neural Networks

use a special kind of linear mathematical operation called convolution. The hidden layers of a CNN typically consist of a series of convolutional layers.[22] CNNs showed great potential for melanoma subtypes and localization diagnosis on dermoscopic image datasets (780 images) and achieved AUROC ≈0.93.[55] Improvements in the accuracy of this model could be achieved by adding more training images of mucosal and subungual sites. Data-augmentation deep models (DADLM) that enhance the learnability of CNNs and Convolutional LSTM (ConvLSTM) deep learning models, improve the accuracy of COVID-19 detection.[56] The study used 50 images (X-ray and CT). This model needs more reliable data to confirm its performance. The fast-track COVID-19 classification network (FCONet) was developed to diagnose COVID-19 pneumonia in CT images (3,993) and differentiate it from non-COVID-19 pneumonia and non-pneumonia diseases with ≈99% accuracy.[57] A CNN was also utilized to classify solid, lipid-poor, contrast enhancing renal masses using enhanced CT images (143 patients) with the accuracy ≈99%, and AUROC ≈0.82,[58] and for automated prediction of breast cancer risk on 92 histopathological images where the F1 score was 0.73.[59] These CNN models are examples of deep learning that rely on medical imaging. More research that uses EHR, or medical text data in addition to imaging could contribute to faster and cheaper diagnostics. Transfer learning is applied to train ResNet-50 and ResNet-101 deep learning models on augmented HAM10000 datasets, which contained about 42,000 dermoscopy skin cancer images. Achieved accuracy was better when used augmented dataset compared to the original dataset and it was about 91.7%.[60] CNN mpdel has shown great success in decoding motor preparation of upper limbs from time-frequency maps of EEG signals.[61] A deep learning architecture was applied to early diagnosis of glaucoma (301 images, AUROC 0.92),[62] and for early diabetic retinopathy detection,[63] on retinal fundus images (40 images, AUROC 0.94). Further studies with larger datasets, adding post-processing methods, and improved optimized deep ML architectures could increase the accuracy of these models. An interpretable classification approach of ultrasound images for the risk assessment and stratification of patients with carotid atheromatous plaque was designed using CNNs and achieved AUROC of 0.73.[64] The integration of interpretability methods with deep learning strategies can facilitate the identification of ultrasound image biomarkers for the stratification of patients with carotid atheromatous plaque. A dataset of 1,900 chest X-ray images has been used with the proposed CNN based model: “C19D-Net” to detect COVID-19 with the accuracy of 96.24%.[65] The idea is to employ the constructed CNN ML model to help radiologists improve their accuracy of detection of COVID-19 from X-ray images.

#### Recurrent Neural Networks

are a type of neural networks that allow analysis of temporal heterogenous medical data. LSTM or GRU units effectively model the irregular visiting patterns in the long, heterogenous sequence of events in EHR.[23] RNNs and the magnetic induction system were integrated to detect a wide range of human motions.[66] The benefit of LSTM RNN for sequence classification is the ability to support multiple parallel temporal input data from different sensor modalities.[66] LSTM and GRU RNN models were developed to predict complications of DM2 (two million patients with DM2 diagnosis), with the prediction accuracy up to 84%. They outperformed traditional ML models in prediction accuracy of 10 selected complications of DM2.[67] An RNN approach was used for predicting hemoglobin levels in patients with end-stage renal disease (7,739 patients) and produced mean absolute error (MAE) of 0.54.[68] Further research is needed to incorporate the dialysis and laboratory information. RNNs were designed for monitoring of depth of anesthesia based on features of EEG signals (20 patients).[69] LSTM RNN models predicted AD from conditions, measurement, and drugs domain on about 2,600 patients. A successful application of the drugs domain in prediction of AD was presented (area under the precision recall curve (AUPRC) 0.99).[70] Additional research with the drugs domain is required to develop comprehensive clinically applicable ML solutions. Another study utilized arterial waveforms recorded on 18,813 patients during noncardiac surgery to predict short-term intraoperative hypotension. A weighted average hybrid of deep learning CNN and RNN models performed the best (AUPRC 0.716).[71] Intraoperative hypotension has an adverse impact on postoperative outcomes and accurate prediction could improve survival. Acute kidney injury (AKI) is associated with poor patient outcomes and increased health care costs. Two RNN algorithms were created using a dataset of more than 72,000 patients.[72] Model 1 predicted the occurrence of AKI within 7 days with AUROC of 0.84 and model 2 predicted the future trajectory of creatinine values up to 72 hours with AUROC of 0.9. Further development of the suggested approaches could incorporate the model into CDS systems for prediction of in-hospital AKI.[72] Researchers leveraged a big dataset (48,151 patients) to build an RNN to predict the risk of developing hepatocellular carcinoma (HCC). RNN models achieved AUROC of 0.759.[73] Deep learning RNN models outperformed traditional models, suggesting that RNN models could be used to identify high risk of developing HCC. The performance of the presented RNN models could be improved by training them on better quality hospital datasets and further optimization of deep learning models. We summarized the reviewed ML models in Table 2.

**Table 2.**
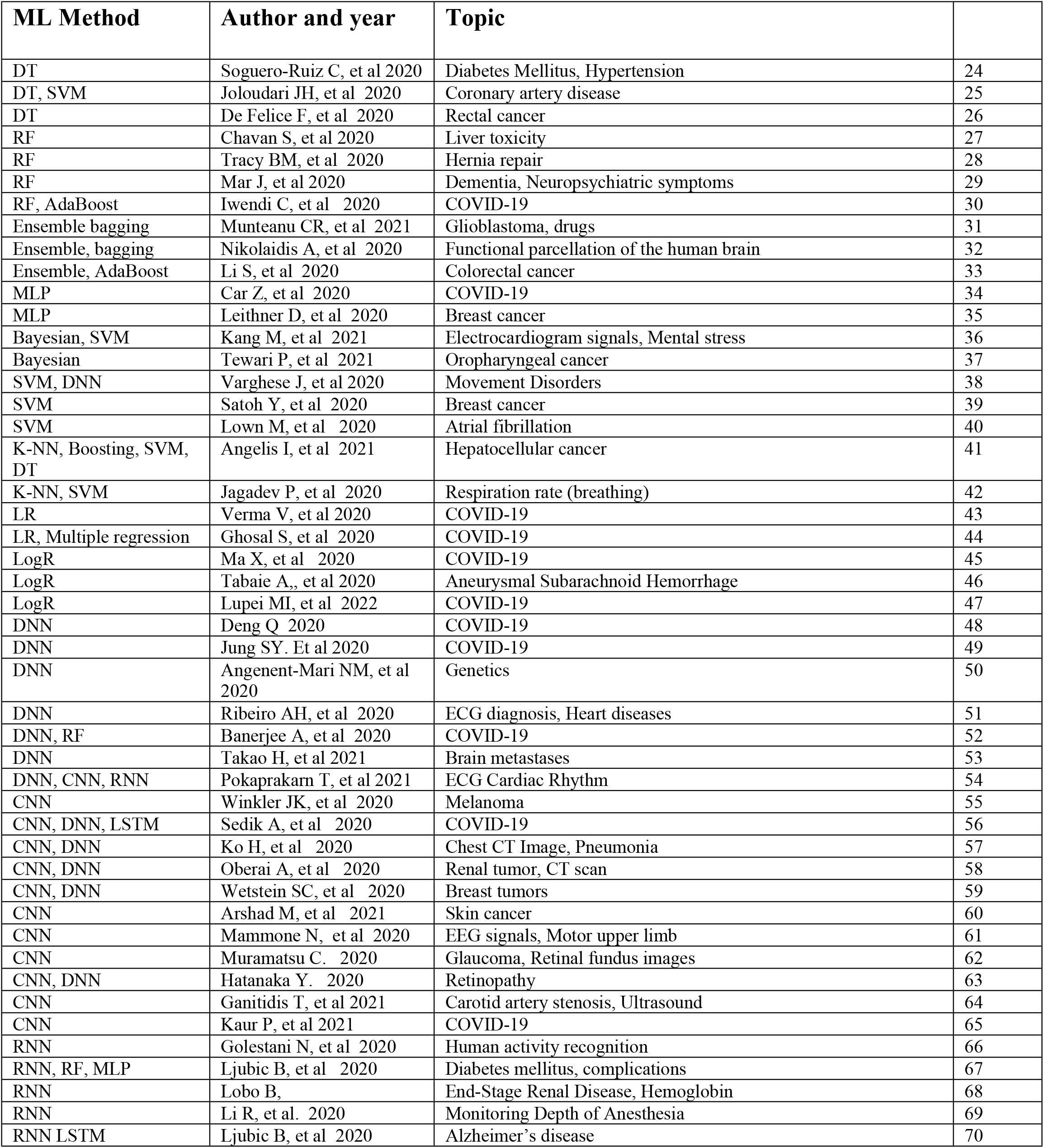
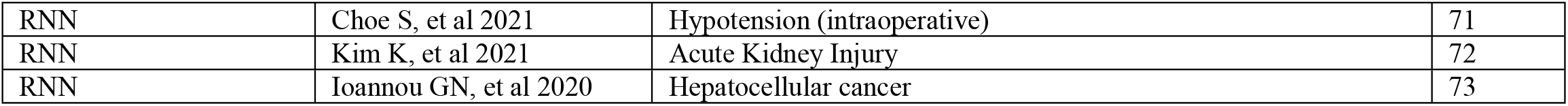
Reviewed articles, classified by ML model types. First authors, publication year, and medical topics described in the publications.

The number of articles by types of medical conditions and topics is presented in Table 3.

**Table 3.** The number of analyzed articles by types of medical conditions.

| Types of diseases by systems or therapy | Number of reviewed articles |
| --- | --- |
| Neoplasms (Oncology) | 13 |
| Nervous System and Sense Organs | 12 |
| COVID-19 | 11 |
| Circulatory System | 10 |
| Endocrine, Nutritional and Metabolic | 2 |
| Respiratory System | 2 |
| Genitourinary System | 2 |
| Mental Disorders | 2 |
| Digestive System | 1 |
| Surgical procedures | 1 |
| Genomics, proteomics | 1 |
| New drugs, drug therapy | 1 |

## DISCUSSION

This review shows that most ML models in medicine represent great software solutions with high prediction accuracy, but only handful of models could find an implementation in medical practice. Neurological conditions are among the most common medical system subject to ML model applications.[29,31,32,38,46,53,61-63,66,69,70] The most frequent type of data used in these applications were imaging data. Images consist of spatially coherent pixels in a local region, meaning that pixels close to each other share similar information. Deep learning architectures (especially CNN) produce higher accuracy predictions from image inputs than from EHR type of datasets, which are often heterogeneous. Another medical discipline extensively used in ML analysis is oncology. Accurate predicting the development of cancers or complications of cancers could indicate earlier diagnosis and therapeutic approaches that would improve outcomes. [26,31,33,35,37,39,41,53,55,58-60,73] The majority of these ML applications use imaging data (most often histologic type) for classification of malignant versus benign tumors. Cardiovascular conditions and DM are among the most common medical conditions used in predictive analysis. [24,25,36,40,46,51,54,64,67,71] The challenges with these types of predictions are often related to limitations of data availability. Insurance claims data was frequently used but often lacks important clinical information such as laboratory results and medications.[67,70]

Many traditional and deep ML models were utilized with the goal of helping to detect COVID-19 infections, complications, or outcomes as one of the most frequent research topic in the last two years. [30,34,43-45,47-49,51,52,56,65]

The performance of predictive ML models in medicine depends on multiple factors. For challenging prediction problems, the understanding of disease is likely to lead to more accurate prediction. Physicians must be better motivated to use ML developments, which is not always easy to achieve since they perceive this activity as something that decreases their time with patients.[1] Since many physicians use computers daily, better presented benefits of ML prediction models could increase their adoption in medicine. Evidence-based medicine requires statistical analysis of medical data and ML is a form of that analysis. Some form of ML should become a part of statistics teaching in medical school to prepare future physicians for meaningful adoption of medical ML models. To make ML more meaningful in clinical practice, we should focus on tasks that physicians need help with and where results of ML could help physicians to improve their decisions. The computers that physicians use for EHR could also be used for ML models. Additionally, ML is relatively inexpensive compared to basic science and large-scale clinical research.

Traditional ML methods do not always achieve accuracy that would convince medical doctors of the benefits of the proposed predictive models.[24-47] A prediction accuracy of 70-90% is generally a good result in terms of performances of ML models but may not be high enough to suggest clinically meaningful improvements in practice. Traditional models have the advantage of simplicity and interpretability but suffer from somewhat worse accuracy.[38,52,67]

The most successful and meaningful application of deep learning ML models was achieved in the imaging field.[53,55-65] Analyses of CT scans, X-rays, Doppler ultrasound, histo-pathological images obtained high accuracy results, which often outperform medical experts. RNN models capture the temporal nature of EHR, imaging and other medical data to predict diseases, complications, and outcomes.[66-73] Deep learning models produce higher accuracy but suffer from issues of interpretability and instability.[15,75] Combinations of traditional and deep learning models could address challenges of interpretability and accuracy.[38,52,67] Many datasets are small and do not have enough samples for implementation of deep learning models. In those cases, traditional ML models are the only option.

To build effective ML models, we must understand how to select relevant features to train ML models. Computational methods that use optimization function to automatically select useful features have been developed.[76,77] In addition to automatically selected features, we often use medical domain knowledge to identify useful features that could help in improvement of predictions.[67,70] If analyses point toward certain features as the most important for obtaining the model performance, the next challenge is how to quantify the relevance of those features.

An important dilemma is the selection of the most optimal ML models for a specific task. We need to consider which specific medical problem we want to solve. We must also determine how much and what type of data are available, how much data are missing, and whether temporal information is included. Implementation of ML in prediction of medical conditions using EHRs and other non-imaging data as cheaper source of data could achieve meaningful results at lower cost. We would need to weigh positive and negative factors in each of the options before we select the most optimal model for the given task. Predictive ML models could potentially help to build CDS systems to make better medical decisions. These models can provide recommendations to select suitable patients for clinical trials.[70] Domain knowledge and collaborations between physicians and ML experts can improve the prediction performance of ML models in medicine and facilitate implementation in clinical practice. Prediction ML models could help clinicians to select the most effective diagnostic and therapeutic choices available. Successful ML models can make medicine more efficient, improve outcomes, and decrease medical errors. We predict that ML models will continue to develop, and they will be applied more broadly in clinical practice.

## Data Availability

All data used in the manuscript are provided as part of the submitted article. Data are extracted from searching the PubMed, publicly available database.

## ACKNOWLEDGMENTS

This research was supported by the National Institute of Diabetes and Digestive and Kidney Diseases of the National Institutes of Health under Award Number R01DK122073. The content is solely the responsibility of the authors and does not necessarily represent the official views of the National Institutes of Health. The research was also supported by Clinical and Translational Science Award (CTSA) Program grants under Award numbers: UL1TR003017, KL2TR003018 and TL1TR003019.

## AUTHORS CONTRIBUTORSHIP

BL, MP, AG, DR, GC, and Z.O. performed the literature review and wrote the paper. The authors have no competing interests to declare.

## REFERENCES

1. Hoyt RE, Yoshihashi AK. Health Informatics: Practical Guide for Healthcare and Information Technology Professionals, Sixth Edition, Morrisville, PA: Lulu Press 2014.

2. Gligorijevic D, Stojanovic J, Djuric N, et al. Large-scale discovery of disease-disease and disease-gene associations. Scientific reports 2016;6(1):1–2.

3. Gligorijevic D, Stojanovic J, Satz W, et al. Deep attention model for triage of emergency department patients. In Proceedings of the 2018 SIAM International Conference on Data Mining, pp 297–305, 2018.

4. Stojanovic J, Gligorijevic D, Radosavljevic V, et al. Modeling healthcare quality via compact representations of electronic health records. IEEE/ACM transactions on computational biology and bioinformatics 2016;14(3):545–54.

5. Waringa J, Lindvall C, Umeton R. Automated machine learning: Review of the state-of-the-art and opportunities for healthcare. https://doi.org/10.1016/j.artmed.2020.101822.

6. Hamet P, Tremblay J. Artificial intelligence in medicine. Metabolism 2017;69:36–40. https://doi.org/10.1016/j.metabol.2017.01.011.

7. Daume H. A Course in Machine Learning. Second edition. 2017. http://ciml.info/dl/v0_99/ciml-v0_99-all.pdf.

8. Breiman L, Friedman J, Stone CJ, et al. Classification and regression trees. Boca Raton, FL: CRC press 1984.

9. Breiman L. Random forests. Machine learning 2001;45(1):5–32.

10. Breiman L. Bagging predictors. Machine learning 1996;24(2):123–40.

11. Schapire RE, Freund Y. Boosting: Foundations and algorithms. Cambridge, MA: MIT Press 2014.

12. Arsov N, Pavlovski M, Basnarkov L, et al. Generating highly accurate prediction hypotheses through collaborative ensemble learning. Scientific reports 2017;7:44649.

13. Pavlovski M, Zhou F, Stojkovic I, et al. Adaptive skip-train structured regression for temporal networks. In Joint European Conference on Machine Learning and Knowledge Discovery in Databases 2017 Sep 18, pp. 305–21. Springer, Cham.

14. Pavlovski M, Zhou F, Arsov N, et al. Generalization-Aware Structured Regression towards Balancing Bias and Variance. In IJCAI 2018 Jul 13, pp. 2616–22.

15. The perceptron. A Course in Machine Learning. Second edition. 2017. http://ciml.info/dl/v0_99/ciml-v0_99-ch04.pdf

16. Rumelhart DE, Geoffrey EH, Williams RJ. “Learning Internal Representations by ErrorPropagation”. David E. Rumelhart, James L. McClelland, and the PDP research group. (editors), Parallel distributed processing: Explorations in the microstructure of cognition, Volume 1: Foundation. Cambridge, MA: MIT Press 1986.

17. Barber D. Bayesian reasoning and machine learning. Cambridge, MA: Cambridge University Press 2012.

18. Cortes C, Vapnik V. Support-vector networks. Machine learning 1995;20(3):273–97.

19. Cover T, Hart P. Nearest neighbor pattern classification[J]. Information Theory, IEEE Trans Inf Theory 1967;13(1):21–7.

20. Weisberg S. Applied linear regression. Hoboken, NJ: John Wiley & Sons 2005

21. Murphy K. Logistic regression. Machine Learning: A Probabilistic Perspective, Chapter 8, pp. 245 –279. Cambridge, MA: MIT Press 2012.

22. Goodfellow I, Bengio Y, Courville A. Deep learning. Cambridge, MA: MIT press 2016.

23. Williams RJ, Zipser D. A learning algorithm for continually running fully recurrent neural networks. Neural Comput 1989;1(2):270–80.

24. Soguero-Ruiz C, Mora-Jiménez I, Mohedano-Munoz MA, et al. Visually guided classification trees for analyzing chronic patients. BMC Bioinformatics 2020;21(Suppl 2):92. doi:10.1186/s12859-020-3359-3

25. Joloudari JH, Joloudari EH, Saadatfar H, et al. Coronary Artery Disease Diagnosis; Ranking the Significant Features Using a Random Trees Model. Int J Environ Res Public Health 2020;17(3):731. doi: 10.3390/ijerph17030731.

26. De Felice F, Crocetti D, Parisi M, et al. Decision tree algorithm in locally advanced rectal cancer: an example of over-interpretation and misuse of a machine learning approach. J Cancer Res Clin Oncol 2020;146(3):761–5. doi:10.1007/s00432-019-03102-y

27. Chavan S, Scherbak N, Engwall M, et al. Predicting Chemical-Induced Liver Toxicity Using High-Content Imaging Phenotypes and Chemical Descriptors: A Random Forest Approach. Chem Res Toxicol 2020;33(9):2261–75. doi:10.1021/acs.chemrestox.9b00459

28. Tracy BM, Finnegan TM, Smith RN, et al. Random forest modeling using socioeconomic distress predicts hernia repair approach. Surg Endosc 2020. doi:10.1007/s00464-020-07860-6

29. Mar J, Gorostiza A, Ibarrondo O, et al. Validation of Random Forest Machine Learning Models to Predict Dementia-Related Neuropsychiatric Symptoms in Real-World Data. J Alzheimers Dis 2020;77(2):855–64. doi:10.3233/JAD-200345

30. Iwendi C, Bashir AK, Peshkar A, et al. COVID-19 Patient Health Prediction Using Boosted Random Forest Algorithm. Front Public Health 2020;8:357. doi:10.3389/fpubh.2020.00357

31. Munteanu CR, Gutiérrez-Asorey P, Blanes-Rodríguez M, et al. Prediction of Anti-Glioblastoma Drug-Decorated Nanoparticle Delivery Systems Using Molecular Descriptors and Machine Learning. Int J Mol Sci 2021;22(21):11519. doi: 10.3390/ijms222111519.

32. Nikolaidis A, Solon Heinsfeld A, Xu T, et al. Bagging improves reproducibility of functional parcellation of the human brain. Neuroimage 2020;214:116678. doi:10.1016/j.neuroimage.2020.116678

33. Li S, Zeng Y, Chapman WC Jr, et al. Adaptive Boosting (AdaBoost)-based multiwavelength spatial frequency domain imaging and characterization for ex vivo human colorectal tissue assessment. J Biophotonics 2020;13(6):e201960241. doi:10.1002/jbio.201960241

34. Car Z, Baressi Šegota S, Anđelić N, et al. Modeling the Spread of COVID-19 Infection Using a Multilayer Perceptron. Comput Math Methods Med 2020;2020:5714714. doi:10.1155/2020/5714714

35. Leithner D, Mayerhoefer ME, Martinez DF, et al. Non-Invasive Assessment of Breast Cancer Molecular Subtypes with Multiparametric Magnetic Resonance Imaging Radiomics. J Clin Med 2020;9(6):1853. doi:10.3390/jcm9061853

36. Kang M, Shin S, Zhang G, et al. Mental Stress Classification Based on a Support Vector Machine and Naive Bayes Using Electrocardiogram Signals. Sensors (Basel) 2021;21(23):7916. doi: 10.3390/s21237916.

37. Tewari P, Kashdan E, Walsh C, et al. Estimating the conditional probability of developing human papilloma virus related oropharyngeal cancer by combining machine learning and inverse Bayesian modelling. PLoS Comput Biol. 2021;17(8):e1009289. doi: 10.1371/journal.pcbi.1009289.

38. Varghese J, Fujarski M, Hahn T, et al. The Smart Device System for Movement Disorders: Preliminary Evaluation of Diagnostic Accuracy in a Prospective Study. Stud Health Technol Inform 2020;270:889–93. doi:10.3233/SHTI200289

39. Satoh Y, Tamada D, Omiya Y, et al. Diagnostic Performance of the Support Vector Machine Model for Breast Cancer on Ring-Shaped Dedicated Breast Positron Emission Tomography Images. J Comput Assist Tomogr 2020;44(3):413–8. doi:10.1097/RCT.0000000000001020

40. Lown M, Brown M, Brown C, et al. Machine learning detection of Atrial Fibrillation using wearable technology. PLoS One 2020;15(1):e0227401. doi:10.1371/journal.pone.0227401

41. Angelis I, Exarchos T. Hepatocellular Carcinoma Detection Using Machine Learning Techniques. Adv Exp Med Biol 2021;1338:21–29. doi: 10.1007/978-3-030-78775-2_4.

42. Jagadev P, Giri LI. Human respiration monitoring using infrared thermography and artificial intelligence. Biomed Phys Eng Express 2020;6(3):035007. doi: 10.1088/2057-1976/ab7a54.

43. Verma V, Vishwakarma RK, Verma A, et al. Time-to-Death approach in revealing Chronicity and Severity of COVID-19 across the World. PLoS One 2020;15(5):e0233074. doi:10.1371/journal.pone.0233074

44. Ghosal S, Sengupta S, Majumder M, et al. Linear Regression Analysis to predict the number of deaths in India due to SARS-CoV-2 at 6 weeks from day 0 (100 cases -March 14th 2020). Diabetes Metab Syndr 2020;14(4):311–5. doi:10.1016/j.dsx.2020.03.017

45. Ma X, Ng M, Xu S, et al. Development and validation of prognosis model of mortality risk in patients with COVID-19. Epidemiol Infect 2020;148:e168. doi:10.1017/S0950268820001727

46. Tabaie A, Nemati S, Allen JW, et al. Assessing Contribution of Higher Order Clinical Risk Factors to Prediction of Outcome in Aneurysmal Subarachnoid Hemorrhage Patients. AMIA Annu Symp Proc 2020;2019:848–56.

47. Lupei MI, Li D, Ingraham NE, et al. A 12-hospital prospective evaluation of a clinical decision support prognostic algorithm based on logistic regression as a form of machine learning to facilitate decision making for patients with suspected COVID-19. PLoS One 2022;17(1):e0262193. doi: 10.1371/journal.pone.0262193.

48. Deng Q. Dynamics and Development of the COVID-19 Epidemic in the United States: A Compartmental Model Enhanced with Deep Learning Techniques. J Med Internet Res 2020;22(8):e21173. doi: 10.2196/21173

49. Jung SY, Jo H, Son H, et al. Real-World Implications of a Rapidly Responsive COVID-19 Spread Model with Time-Dependent Parameters via Deep Learning: Model Development and Validation. J Med Internet Res 2020;22(9):e19907. doi: 10.2196/19907

50. Angenent-Mari NM, Garruss AS, Soenksen LR, et al. A deep learning approach to programmable RNA switches. Nat Commun 2020;11(1):5057. doi:10.1038/s41467-020-18677-1

51. Ribeiro AH, Ribeiro MH, Paixão GMM, et al. Automatic diagnosis of the 12-lead ECG using a deep neural network. Nat Commun 2020;11(1):1760. doi:10.1038/s41467-020-15432-4

52. Banerjee A, Ray S, Vorselaars B, et al. Use of Machine Learning and Artificial Intelligence to predict SARS-CoV-2 infection from Full Blood Counts in a population. Int Immunopharmacol 2020;86:106705. doi:10.1016/j.intimp.2020.106705

53. Takao H, Amemiya S, Kato S, et al. Deep-learning single-shot detector for automatic detection of brain metastases with the combined use of contrast-enhanced and non-enhanced computed tomography images. Eur J Radiol 2021;144:110015. doi: 10.1016/j.ejrad.2021.110015.

54. Pokaprakarn T, Kitzmiller RR, Moorman R, et al. Sequence to Sequence ECG Cardiac Rhythm Classification using Convolutional Recurrent Neural Networks. IEEE J Biomed Health Inform 2021; doi: 10.1109/JBHI.2021.3098662.

55. Winkler JK, Sies K, Fink C, et al. Melanoma recognition by a deep learning convolutional neural network-Performance in different melanoma subtypes and localisations. Eur J Cancer 2020;127:21–9. doi:10.1016/j.ejca.2019.11.020

56. Sedik A, Iliyasu AM, Abd El-Rahiem B, et al. Deploying Machine and Deep Learning Models for Efficient Data-Augmented Detection of COVID-19 Infections. Viruses 2020;12(7):769. doi: 10.3390/v12070769

57. Ko H, Chung H, Kang WS, et al. COVID-19 Pneumonia Diagnosis Using a Simple 2D Deep Learning Framework with a Single Chest CT Image: Model Development and Validation. J Med Internet Res 2020;22(6):e19569. doi:10.2196/19569

58. Oberai A, Varghese B, Cen S, et al. Deep learning based classification of solid lipid-poor contrast enhancing renal masses using contrast enhanced CT. Br J Radiol 2020;93(1111):20200002. doi:10.1259/bjr.20200002

59. Wetstein SC, Onken AM, Luffman C, et al. Deep learning assessment of breast terminal duct lobular unit involution: Towards automated prediction of breast cancer risk. PLoS One 2020;15(4):e0231653. doi:10.1371/journal.pone.0231653

60. Arshad M, Khan MA, Tariq U, et al. A Computer-Aided Diagnosis System Using Deep Learning for Multiclass Skin Lesion Classification. Comput Intell Neurosci 2021;2021:9619079. doi:10.1155/2021/9619079

61. Mammone N, Ieracitano C, Morabito FC. A deep CNN approach to decode motor preparation of upper limbs from time-frequency maps of EEG signals at source level. Neural Netw 2020;124:357–72. doi:10.1016/j.neunet.2020.01.027

62. Muramatsu C. Diagnosis of Glaucoma on Retinal Fundus Images Using Deep Learning: Detection of Nerve Fiber Layer Defect and Optic Disc Analysis. Adv Exp Med Biol 2020;1213:121–32. doi:10.1007/978-3-030-33128-3_8

63. Hatanaka Y. Retinopathy Analysis Based on Deep Convolution Neural Network. Adv Exp Med Biol 2020;1213:107–20. doi: 10.1007/978-3-030-33128-3_7

64. Ganitidis T, Athanasiou M, Dalakleidi K, et al. Stratification of carotid atheromatous plaque using interpretable deep learning methods on B-mode ultrasound images. Annu Int Conf IEEE Eng Med Biol Soc 2021;2021:3902–3905. doi: 10.1109/EMBC46164.2021.9630402.

65. Kaur P, Harnal S, Tiwari R, et al. A Hybrid Convolutional Neural Network Model for Diagnosis of COVID-19 Using Chest X-ray Images. Int J Environ Res Public Health 2021;18(22):12191. doi: 10.3390/ijerph182212191.

66. Golestani N, Moghaddam M. Human activity recognition using magnetic induction-based motion signals and deep recurrent neural networks. Nat Commun 2020;11(1):1551. doi: 10.1038/s41467-020-15086-2

67. Ljubic B, Abdel Hai A, Stanojevic M, et al. Predicting Complications of Diabetes Mellitus Using Advanced Machine Learning Algorithms. JAMIA 2020;27(9):1343–51. doi:10.1093/jamia/ocaa120

68. Lobo B, Abdel-Rahman E, Brown D, et al. A recurrent neural network approach to predicting hemoglobin trajectories in patients with End-Stage Renal Disease. Artif Intell Med 2020;104:101823. doi:10.1016/j.artmed.2020.101823

69. Li R, Wu Q, Liu J, et al. Monitoring Depth of Anesthesia Based on Hybrid Features and Recurrent Neural Network. Front Neurosci 2020;14:26. doi:10.3389/fnins.2020.00026

70. Ljubic B, Roychoudhury S, Cao XH, et al. Influence of medical domain knowledge on deep learning for Alzheimer’s disease prediction. Comput Methods Programs Biomed 2020;197:105765. doi:10.1016/j.cmpb.2020.105765

71. Choe S, Park E, Shin W, et al. Short-Term Event Prediction in the Operating Room (STEP-OP) of Five-Minute Intraoperative Hypotension Using Hybrid Deep Learning: Retrospective Observational Study and Model Development. JMIR Med Inform 2021;9(9):e31311. doi: 10.2196/31311.

72. Kim K, Yang H, Yi J, Son HE, et al. Real-Time Clinical Decision Support Based on Recurrent Neural Networks for In-Hospital Acute Kidney Injury: External Validation and Model Interpretation. J Med Internet Res 2021;23(4):e24120. doi: 10.2196/24120.

73. Ioannou GN, Tang W, Beste LA, et al. Assessment of a Deep Learning Model to Predict Hepatocellular Carcinoma in Patients with Hepatitis C Cirrhosis. JAMA Netw Open 2020;3(9):e2015626. doi: 10.1001/jamanetworkopen.2020.15626.

74. https://www.hcup-us.ahrq.gov/databases.jsp.

75. Antun V, Renna F, Poon C, et al. On instability of deep learning in image reconstruction and the potential costs of AI. Proceedings of the National Academy of Sciences of the United States of America 2020;117(48):30088–95

76. Stojkovic I, Jelisavcic V, Milutinovic V, et al. Distance Based Modeling of Interactions in Structured Regression. In IJCAI 2016;2032–8.

77. Stojkovic I, Jelisavcic V, Milutinovic V, et al. Fast Sparse Gaussian Markov Random Fields Learning Based on Cholesky Factorization. In IJCAI 2017;2758–64.

